# Increasing test specificity without impairing sensitivity – lessons learned from SARS-CoV-2 serology

**DOI:** 10.1101/2020.11.05.20226449

**Authors:** Thomas Perkmann, Thomas Koller, Nicole Perkmann-Nagele, Maria Oszvar-Kozma, David W Eyre, Philippa Matthews, Abbie Bown, Nicole Stoesser, Marie-Kathrin Breyer, Robab Breyer-Kohansal, Otto C Burghuber, Sylvia Hartl, Daniel Aletaha, Daniela Sieghart, Peter Quehenberger, Rodrig Marculescu, Patrick Mucher, Astrid Radakovics, Miriam Klausberger, Mark Duerkop, Barbara Holzer, Boris Hartmann, Robert Strassl, Gerda Leitner, Florian Grebien, Wilhelm Gerner, Reingard Grabherr, Oswald F Wagner, Christoph J Binder, Helmuth Haslacher

## Abstract

**Background:** Serological tests are widely used in various medical disciplines for diagnostic and monitoring purposes. Unfortunately, the sensitivity and specificity of test systems is often poor, leaving room for false positive and false negative results. However, conventional methods used to increase specificity decrease sensitivity and vice versa. Using SARS-CoV-2 serology as an example, we propose here a novel testing strategy: the “Sensitivity Improved Two-Test” or “ SIT^2^” algorithm.

**Methods:** SIT^2^ involves confirmatory re-testing of samples with results falling in a predefined retesting-zone of an initial screening test, with adjusted cut-offs to increase sensitivity. We verified and compared the performance of SIT^2^ to single tests and orthogonal testing (OTA) in an Austrian cohort (1,117 negative, 64 post-COVID positive samples) and validated the algorithm in an independent British cohort (976 negatives, 536 positives).

**Results:** The specificity of SIT^2^ was superior to single tests and non-inferior to OTA. The sensitivity was maintained or even improved using SIT^2^ when compared to single tests or OTA. SIT^2^ allowed correct identification of infected individuals even when a live virus neutralization assay could not detect antibodies. Compared to single testing or OTA, SIT^2^ significantly reduced total test errors to 0.46% (0.24-0.65) or 1.60% (0.94-2.38) at both 5% or 20% seroprevalence.

**Conclusion:** For SARS-CoV-2 serology, SIT^2^ proved to be the best diagnostic choice at both 5% and 20% seroprevalence in all tested scenarios. It is an easy to apply algorithm and can potentially be helpful for the serology of other infectious diseases.

## Introduction

Serological tests are commonly used diagnostic tools in a broad medical field, spanning from infectiology (1) to autoimmunity (2), oncology (3) and transplantation medicine (4). They also play a critical role in animal disease surveillance (5). However, many serological tests come with acceptable but imperfect sensitivities and specificities. Tests with specificities slightly above 90% are considered good (6) or even highly specific (3). However, at low seroprevalence rates, every single percent counts: if the frequency of a given disease in the tested population is only 5%, a specificity of 90% would mean that - even at a sensitivity of 100% - 5 true positives would be matched by ten false positives. Thus, the probability of an individual with a positive test (positive predictive value, PPV) to be a true positive would be only 33%.

During the early phase of the SARS-CoV-2 pandemic, seroprevalences were far below 1% (7). Therefore, highly specific test systems were necessary (>99,5%) to provide good positive predictive values (8). Sensitivity and specificity can be seen as communicating vessels – the improvement of one is usually at the expense of the other (9). Consequently, test systems adjusted by the manufacturers to very high specificities (>99%) showed moderate sensitivity. This problem was particularly evident when non-hospitalized patients were included in the cohorts studied (10-12). To further increase specificity at very low seroprevalence levels, various methods have been proposed, e.g., raising the thresholds for positivity or confirming a positive result with a second test (orthogonal testing) (9, 13, 14). Unfortunately, these specificity improvement strategies inevitably lead to a further reduction of the previously moderate sensitivities.

As the pandemic progressed, the problem became more pronounced as antibodies declined, and sophisticated statistical models were required to compensate for the waning sensitivity (15). In the case of SARS-CoV-2, as with any evolving pandemic, increasing seroprevalence rates worldwide have attenuated the need for higher specificity.

However, the problem persists in non-epidemic diseases where seroprevalence remains low. Moreover, each new pandemic begins with extremely low seroprevalence rates as well, and in the future, we should have better diagnostic strategies in infectious serology ready here.

In the present work, we propose for the first time an orthogonal test algorithm based on real-life data for the SARS-CoV-2 antibody tests of Roche, Abbott, DiaSorin, and two commercial SARS-CoV-2 ELISAs (16) intending to maximize both specificity and sensitivity at the same time. Although our algorithm follows a general principle, it was developed based on SARS-CoV-2 antibody tests. The SARS-CoV-2 pandemic provided a unique opportunity in this regard, as historical samples from before the pandemic are negative by definition (thus allowing accurate specificity testing). In addition, sufficient PCR-confirmed positive cases were available quickly, ensuring a sophisticated and reliable sensitivity verification. Thus, for SARS-CoV-2 - in contrast to most other circulating microorganisms - a realistic and unusually accurate estimation of specificities and sensitivities of serological tests was possible. We benefited from this advantage to develop our sophisticated diagnostic algorithm.

## Methods

### Study design and cohorts

Sera used in this non-blinded prospective cross-sectional study were either residual clinical specimens or samples stored in the MedUni Wien Biobank (n=1,181), a facility specialized in the preservation and storage of human biomaterial, which operates within a certified quality management system (ISO 9001:2015) (17).

For derivation of the SIT^2^ algorithm, sample sets from individuals known to be negative and positive were established for testing. As previously described (18), samples collected before 01.01.2020 (i.e., assumed SARS-CoV-2 negative) were used as a specificity cohort (n=1117): a cross-section of the Viennese population (the LEAD study)(19), preselected for samples collected between November and April to enrich for seasonal infections (n=494); a collection of healthy voluntary donors (n= 265); a disease-specific collection of samples from patients with rheumatic diseases (n=358); (see also Tables S1 and S2).

The SARS-CoV-2 positive cohort (n=64 samples from n=64 individuals) included patients testing positive with RT-PCR during the first wave and their close, symptomatic contacts. Of this cohort five individuals were asymptomatic, 42 had mild-moderate symptoms, four reported severe symptoms, and 13 were admitted to the Intensive Care Unit (ICU). The timing of symptom onset was determined by a questionnaire for convalescent donors and by reviewing individual health records for patients and was in median 41 [26,25-49] days. For asymptomatic donors (n=5), SARS-CoV-2 RT-PCR confirmation time was used instead (for more details, see Tables S1 and S3). All included participants gave written informed consent to donate their samples for research purposes. The overall evaluation plan conformed with the Declaration of Helsinki as well as relevant regulatory requirements. It was reviewed and approved by the ethics committee of the Medical University of Vienna (1424/2020).

For validation of the SIT^2^ algorithm, we used data from an independent United Kingdom cohort (20), including 1,512 serum/plasma samples (536 PCR confirmed SARS-CoV-2 positive cases; 976 negative cases, collected earlier than 2017).

### Antibody testing

For the derivation analyses, SARS-CoV-2 antibodies were either measured according to the manufacturers’ instructions on three different commercially available automated platforms (Roche Elecsys® SARS-CoV-2 [nuclecapsid total antibody assay, further referred to as Roche NC], Abbott SARS-CoV-2-IgG assay [nucleocapsid IgG assay, Abbott NC], DiaSorin LIASION® SARS-CoV-2 S1/S2 assay [S1/S2 combination antigen IgG assay, DiaSorin S1/S2]) or using 96-well enzyme-linked immunosorbent assays (ELISAs) (Technoclone Technozym® RBD and Technozym® NP) yielding quantitative results(16) (for details see Supplement, Supplemental Methods). The antibody assays used in the validation cohort were Abbott NC, DiaSorin S1/S2, Roche NC, Siemens RBD total antibody, and a novel 384-well trimeric spike protein ELISA (Oxford Immunoassay) (20), resulting in 20 evaluable combinations. All samples from the Austrian SARS-CoV-2-positive cohort also underwent live virus neutralization testing (VNT), and neutralization titers (NT) were calculated, as is described in detail in the Supplemental Methods.

### Sensitivity improved two-test method (SIT^2^)

Our newly developed sensitivity improved two-test (SIT^2^) method consists of the following key components: i) sensitivity improvement by cut-off modification and ii) specificity rescue by a second, confirmatory test (Fig. 1A).

**Fig. 1.**
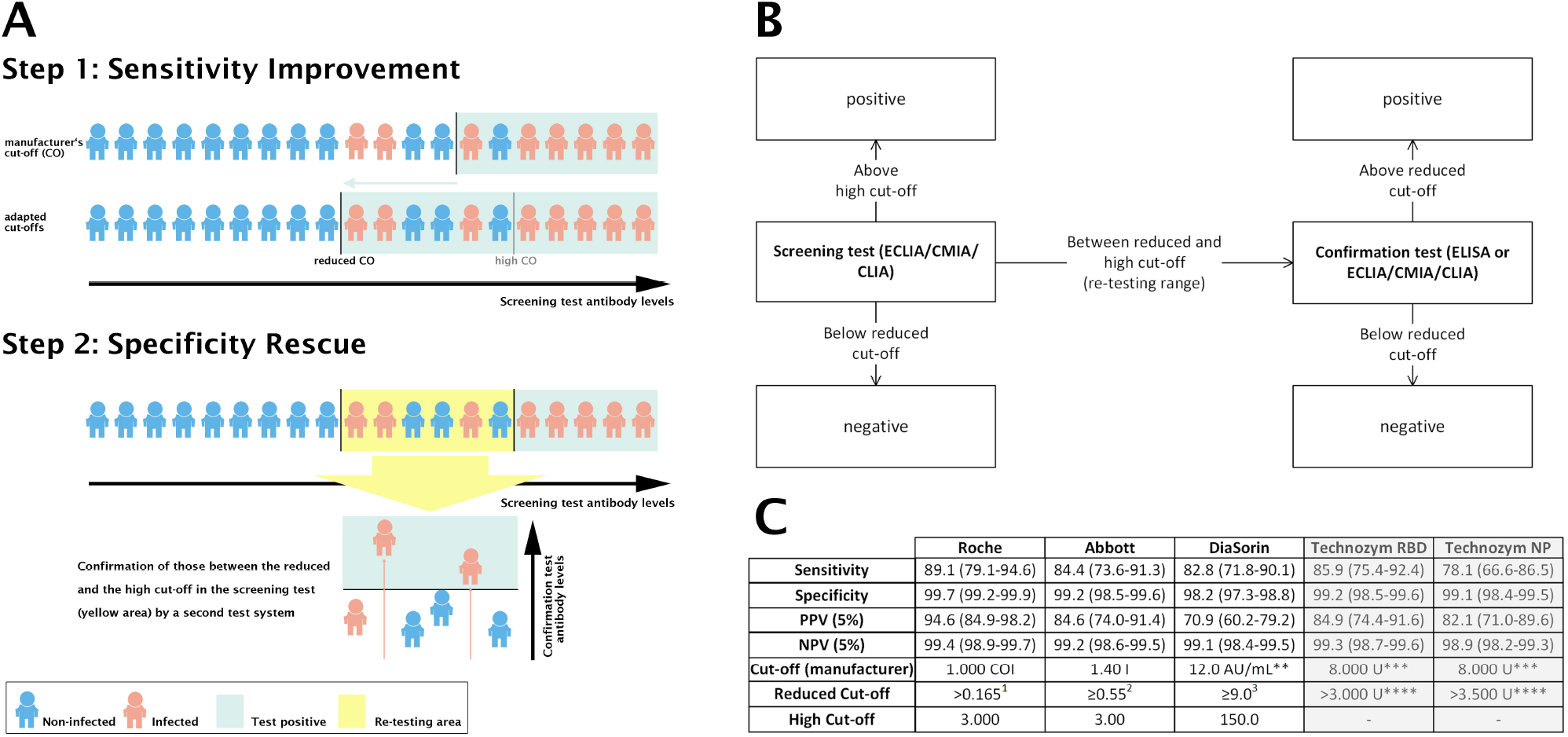
**A)** The Sensitivity Improved Two-Test (SIT^2^) algorithm includes sensitivity improvement by adapted cut-offs and a subsequent specificity rescue by re-testing all samples within the re-testing zone of the screening test by a confirmatory test. **B)** Testing algorithm for SIT^2^ utilizing a screening test on an automated platform (ECLIA/Roche, CMIA/Abbott, CLIA/DiaSorin) and a confirmation test, either on one of the remaining platforms or tested by means of ELISA (Technozym RBD, NP). **C)** All test results between a reduced cut-off suggested by the literature, and a higher cut-off, above which no more false-positives were observed, were subject to confirmation testing. **… results between 12.0 and 15.0, which are according to the manufacturer considered borderline, were treated as positives; ***… suggested as a cut-off for seroprevalence testing; ****… determined by in-house modeling; ^1^… see (21); ^2^… see (22); ^3^… see (23).

For the first component of the SIT^2^ algorithm, positivity thresholds were optimized for sensitivity according to the first published alternative thresholds for the respective assays, calculated e.g. by ROC-analysis (21-23). Additionally, a high cut-off, above which a result can be reliably regarded as true positive without the need for further confirmation, was defined. These levels were based on in-house observations(18) and represent those values (including a safety margin) above which no more false positives were found. The highest results seen in false-positives were 1.800 COI, 2.86 Index, and 104.0 AU/mL, respectively. Hence, we defined the high cut-off for Roche and Abbott as 3.00 COI/Index and for DiaSorin as 150.0 AU/mL. The lowering of positivity thresholds improves sensitivity; the high cut-off prevents unnecessary re-testing of clearly positive samples. Moreover, the high cut-off avoids possible erroneous exclusion by the confirmatory test. The newly defined interval between the reduced threshold for positivity and the high cut-off is the re-testing zone (Fig. 1A). The initial antibody test (screening test) is then followed by a confirmatory test, whereby positive samples from the re-testing zone of the screening test are re-tested. Also, for the confirmatory test, sensitivity-adapted assay thresholds are needed (Figs.1A, 1B). As false-positive samples are usually only positive in one test system (Fig. S1), false positives can be identified, and specificity markedly restored with minimal additional testing as most samples do not fall within the re-testing zone (18, 24). A flowchart of the testing strategy and the applied cut-off levels and their associated quality criteria are presented in Figs. 1B, 1C.

### Test strategy evaluation

On the derivation cohort, we compared the overall performance of the following SARS-CoV-2 antibody testing strategies: i) testing using single assays; ii) simple lowering of thresholds; iii) classical orthogonal testing (OTA), and iv) our newly developed SIT^2^ algorithm at assumed seroprevalences of 5% and 20%. As part of the derivation, we then compared the performance of OTAs and SIT^2^ against the results of a virus neutralization assay. On the validation cohort, we then compared the performance of OTAs and SIT^2^. Finally, we used data from this cohort to evaluate the performance of SIT^2^ versus single tests at seroprevalences of 5%, 10%, 20%, and 50% if the Abbott and DiaSorin assays (i.e., assays with varying degrees of discrepancies in sensitivity and specificity) were used.

### Statistical analysis

Unless otherwise indicated, categorical data are given as counts (percentages), and continuous data are presented as median (interquartile range). Total test errors were compared by Mann-Whitney tests or, in case they were paired, by Wilcoxon tests. 95% confidence intervals (CI) for sensitivities and specificities were calculated according to Wilson, 95% CI for predictive values were computed according to Mercaldo-Wald unless otherwise indicated. Sensitivities and specificities were compared using z-scores. P values <0.05 were considered statistically significant. All calculations were performed using Analyse-it 5.66 (Analyse-it Software, Leeds, UK) and MedCalc 19.6 (MedCalc bvba, Ostend, Belgium). Graphs were drawn using Microsoft Visio (Armonk, USA) and GraphPad Prism 7.0 (La Jolla, USA).

## Results

In the derivation cohort of 1,117 pre-pandemic sera and 64 sera from convalescent COVID-19 patients (80% non-hospitalized, 20% hospitalized), the Roche NC, Abbott NC, and DiaSorin S1/S2 antibody assays gave rise to 7/64, 10/64, and 11/64 false-negative, as well as to 3/1,117, 9/1,117, and 20/1,117 false-positive results. Assuming a seroprevalence of 20%, this led to 2180, 3120, and 3440 false-negative results per 100,000 tests, and 240, 650 and 1,440 false-positive results per 100,000 tests respectively (Fig. 2A, right panel).

**Fig. 2.**
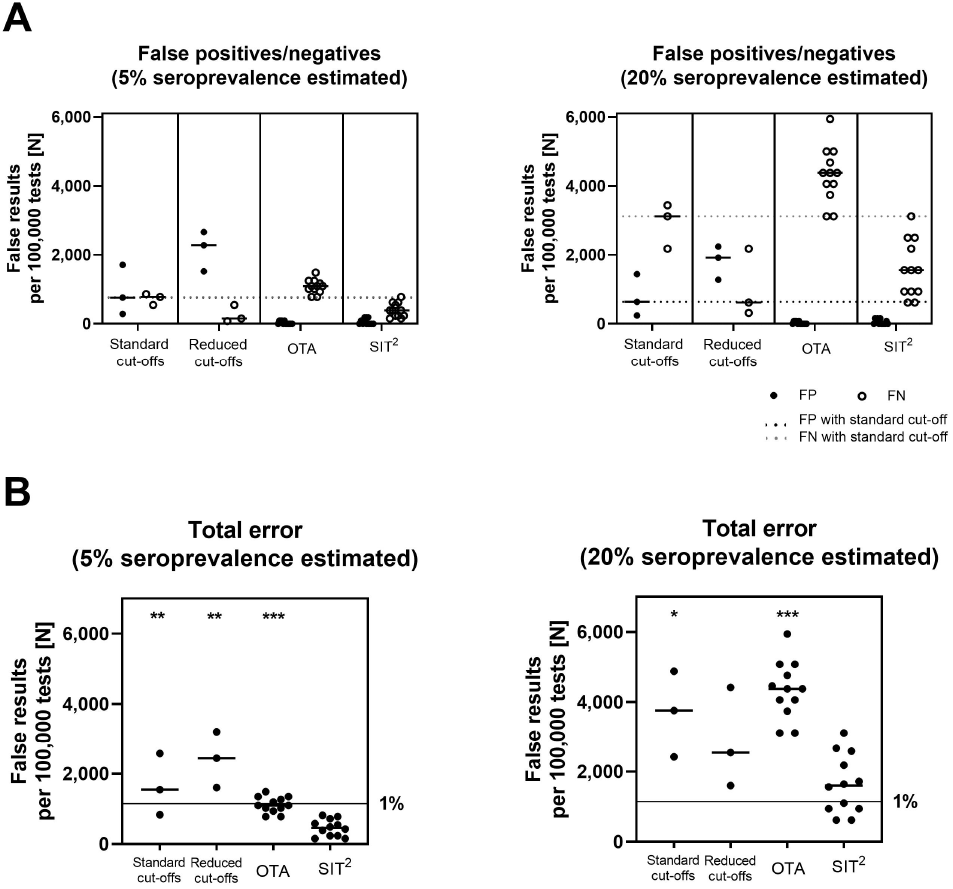
False-positives (FP)/false-negatives (FN) (**A**) and total error (**B**) of single tests, tests with reduced thresholds according to (21-23), orthogonal testing algorithms (OTAs) and the Sensitivity Improved Two-Test (SIT^2^) algorithm at 5 and 20% estimated seroprevalence. Data in (B) were compared by Mann-Whitney tests (unpaired) or Wicoxon tests (paired). *… P<0.05; **…P<0.01; ***…P<0.001.

### Effects of threshold lowering on Sensitivity and Specificity

Lowering the positivity thresholds for the Roche NC, Abbott NC, and Diasorin S1/S2 to 0.165 COI, 0.55 Index and 9 AU/mL increased the sensitivity significantly and reduced false-negative results to 63/64, 62/64, and 57/64 (320, 620, and 2,180 per 100,000 tests at a seroprevalence of 20%), but substantially increased false-positive results to 18/1,117, 27/1,117, and 31/1,117, respectively (1,280, 1,920 and 2,240 per 100,000 tests, an assumed seroprevalence of 20%; Table S4, Fig. 2A, right panel).

### Classical OTA compared to SIT^2^

Subsequently, we evaluated 12 OTA combinations using the fully automated SARS-CoV-2 antibody tests from Roche NC, Abbott NC, and DiaSorin S1/S2 as screening tests, each combined with one of the other fully automated assays or a commercially available NC or RBD-specific ELISA as a confirmation test. Combining these tests as classical OTAs significantly increased specificity and reduced false positives to 0 (0-1)/1,117. However, the rate of false negatives was 14 (12-16)/64 (1,095 [955-1,230] per 100,000 tests at 20% seroprevalence), and therefore considerably higher than for single testing strategies. In contrast, the SIT^2^ algorithm minimized false positives to 0 (0-2)/1,117 (0 [0-140) per 100,000 tests at 20% seroprevalence) while also reducing false negatives to 5 (3-8)/64 (1,560 [940-2420] per 100,000 tests at 20% seroprevalence, Fig. 2A right panel; Table S5).

### Reduction of total error rates by the Sensitivity-Improved Two-Test

Of all the methods assessed, SIT^2^ reached the lowest total error rates per 100,000 tests under both 5% and 20% assumed seroprevalence (455 [235-685] and 1,600 [940-2,490] per 100,000 tests) (Fig. 2B). At a seroprevalence of 5 %, OTA on average performed better than individual tests, and the total error rates of the single tests were higher for the Abbott NC and DiaSorin S1/S2 assay (OTA 1,095 [955-1,325] vs. 830 [Roche NC], 1,540 [Abbott NC] and 2,570 [DiaSorin S1/S2] per 100,000 tests). But with a seroprevalence of 20 %, performance of OTAs, worsened compared to single tests (OTA 4,380 [3,820-5,000] vs 1,600 [Roche], 2,540 [Abbott] and 4,420 [DiaSorin] per 100,000 tests) (Fig. 2B). Therefore, at both 5% and 20% seroprevalence, SIT^2^ resulted in the lowest overall errors. Compared to OTAs, SIT^2^ yielded a similar improvement in specificity while not suffering from the significant sensitivity reduction (Fig. S2). Since the better overall performance of SIT^2^ compared to OTAs was not due to increased specificity but improved sensitivity, we subsequently set out to examine these differences in more detail.

### Sensitivities of single tests, OTA and SIT^2^ in relation to Neutralization Testing

Next, we compared the sensitivities of the three screening tests as single tests and in both two-test methods (OTA and SIT^2^), benchmarking them using the Austrian sensitivity cohort (n=64) simultaneously evaluated with an authentic SARS-CoV-2 virus neutralization test (VNT). Regardless of the screening test used (Roche NC, Abbott NC, or DiaSorin S1/S2), OTAs had lower sensitivities than single tests (80.5% [78.5-83.6], 78.1% [75.8-82.8], or 75.8% [71.5-78.9] vs. 89.1%, 84.4%, or 82.8% respectively), and SIT^2^ showed the best sensitivities of all methods (95.3% [93.0-96.5], 93.8% [92.2-96.5], or 87.5% [85.1-88.7]) (Fig. 3). SIT^2^ algorithms, including the Roche NC and Abbott NC assays, achieved similar or even higher sensitivities than VNT (Fig. 3, VNT reference line), made possible by the unique re-testing zone of SIT^2^ (Fig. S3).

**Fig. 3.**
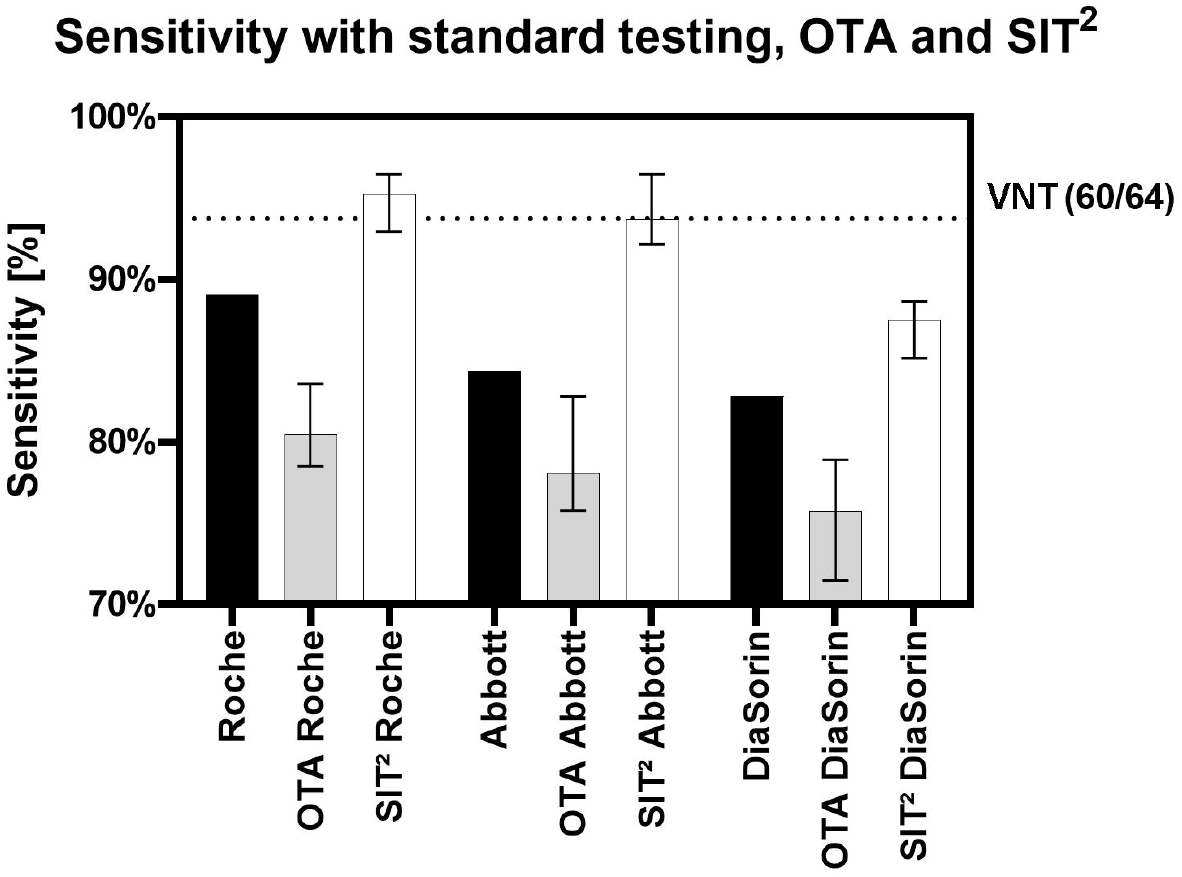
Sensitivities of single tests, orthogonal testing algorithms (OTAs) and the Sensitivity Improved Two-Test (SIT^2^) algorithm. The dotted line indicates the sensitivity of virus neutralization test (VNT).

### Validation of the Sensitivity-Improved Two-Test using an independent cohort

To confirm the improved sensitivity of SIT ^2^ compared to OTA, we analyzed the sensitivities of OTAs and SIT^2^ in an independent validation cohort of 976 pre-pandemic samples and 536 post-COVID samples. Out of 20 combinations using the assays Roche NC (total antibody), Abbott NC (IgG), DiaSorin S1/S2 (IgG), Siemens RBD (total antibody), and Oxford trimeric-S (IgG), a statistically significant improvement in sensitivities over OTAs was shown for SIT^2^ in 18 combinations (Fig. 4). The performance was comparable for the remaining two combinations (Siemens RBD with Oxford trimeric-S and vice versa). Still, no statistically significant improvement could be achieved due to the high pre-existing sensitivities of these assays on this particular sample cohort.

**Fig. 4.**
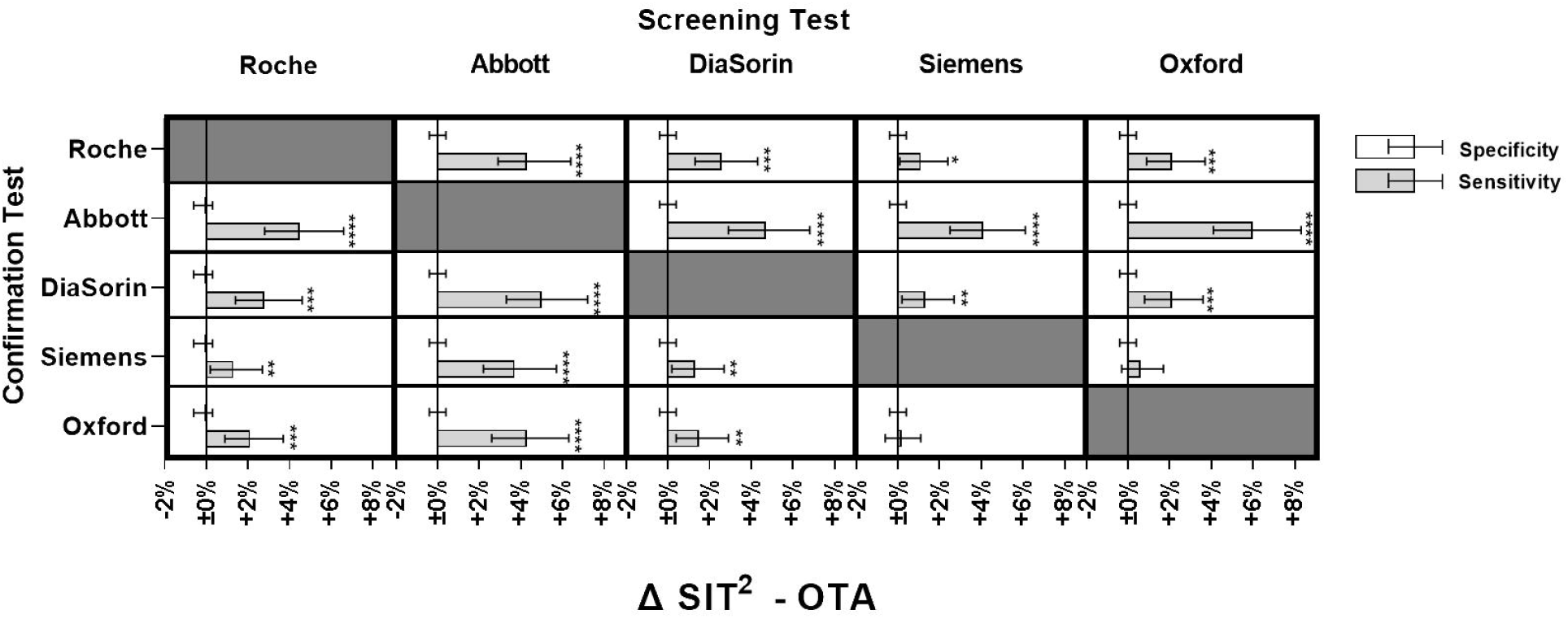
Differences in sensitivity and specificity (mean±95% confidence interval) between the Sensitivity Improved Two-Test (SIT^2^) algorithm and standard orthogonal testing algorithms (OTAs) within the UK validation cohort. *… P<0.05; **…P<0.01; ***…P<0.001; ****…P<0.0001

To further illustrate the effect of SIT^2^ on the outcome of SARS-CoV-2 antibody testing, we compared single testing versus SIT^2^ with the Abbott and DiaSorin assays at varying assumed seroprevalences (5, 10, 20, and 50%), given that the Abbott NC assay is a highly specific (99.9%), but moderately sensitive test (92.7%), and the DiaSorin S1/S2 assay has the most limited specificity (98.7%) of all evaluated assays but an acceptable sensitivity (96.3%). Regardless of whether a lack of specificity (DiaSorin S1/S2) or sensitivity (Abbott NC) had to be compensated for, SIT^2^ improved the overall error rate compared to the individual tests in all four combinations and at all four assumed seroprevalence levels (Fig. 5).

**Fig. 5.**
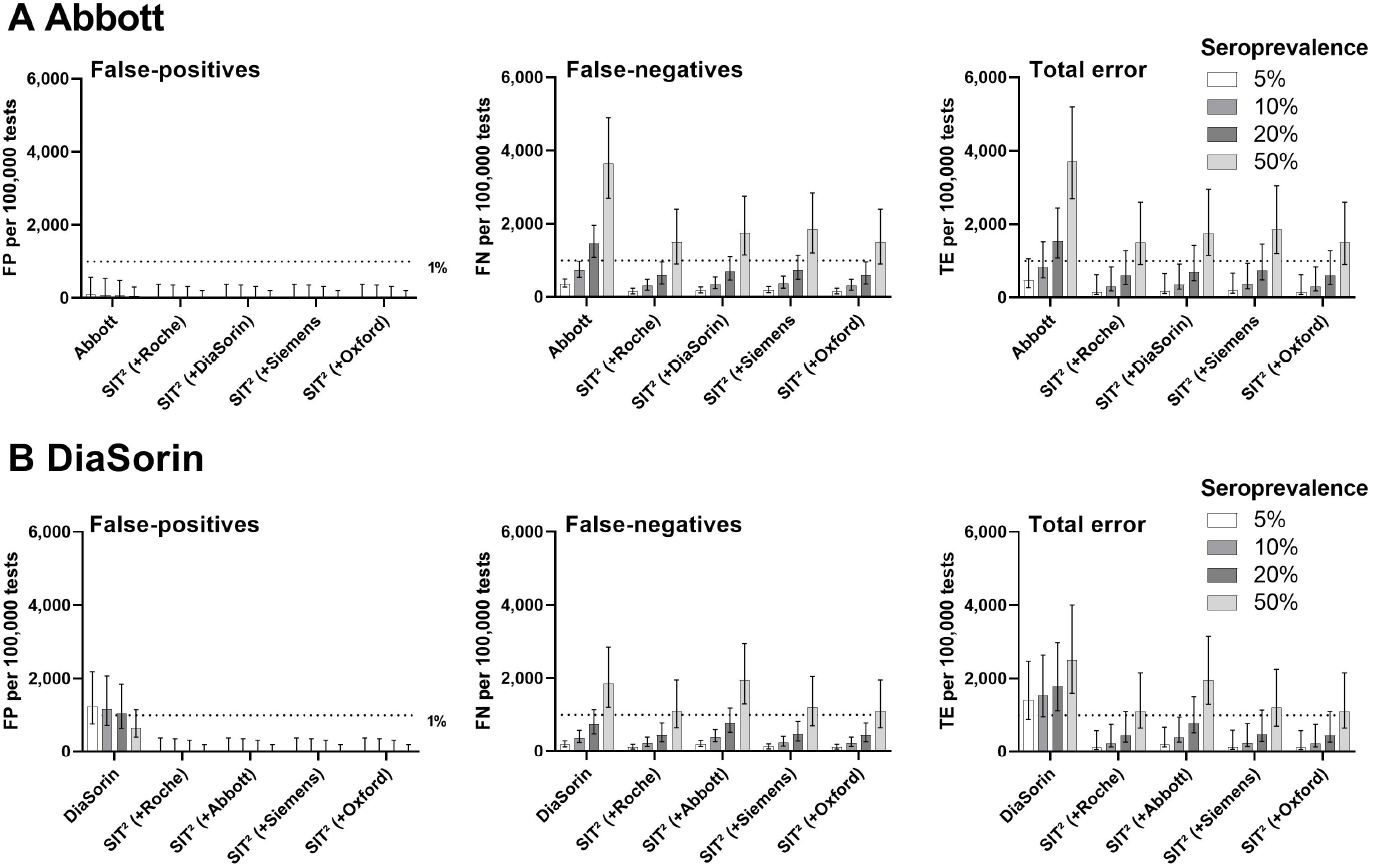
Comparing false-positives (FP), false-negatives (FN), and total error (TE) for two selected test systems, **A)** Abbott, **B)** DiaSorin, between different Sensitivity Improved Two-Test (SIT^2^) combinations and the respective single test within the UK validation cohort for different estimated seroprevalences.

## Discussion

Serology is a commonly used, multi-purpose analytical method (1-4). However, not all serologic assays have appropriate sensitivities and specificities, especially in low-prevalence settings. The SARS-CoV-2 pandemic prompted the simultaneous development of several antibody tests and, which is rare otherwise, allowed to evaluate these tests with both confirmed positive and negative cases, the latter derived from biobank collections established before the virus emerged. In the case of SARS-CoV-2, false-positive samples are usually not simultaneously reactive in different test systems (14, 25). This led to the hypothesis that reducing the threshold for positivity in screening and confirmation tests would increase the specificity without impairing the sensitivity. A further improvement in sensitivity was possible by defining a high cut-off for the screening test, above which, due to the excellent reliability of high test results, no further confirmation (and, thereby, a possible false-negative result in the confirmation test) was necessary.

In the early waves of the SARS-CoV-2 pandemic, many commercially available SARS-CoV-2 antibody tests did not provide sufficient specificity to achieve acceptable positive predictive values (PPVs), for example, at a seroprevalence rate of 1-5% (13, 25). Lowering positivity thresholds might improve test sensitivity (21-23) and conventional orthogonal testing can maximize specificity (26-28). The latter might increase the positive predictive value, but PPV will only be relevantly increased at low seroprevalences. However, since seroprevalence is often neither known and varies widely from region to region, it is difficult to judge whether a less specific or less sensitive test is the lesser of two evils.

However, the reported imperfect specificities and sensitivities of SARS-CoV-2 antibody assays are still considerably higher than those of many other serological tests used in clinical routine. Based on actual data related to SARS-CoV-2, which are in principle generalizable for all types of serological tests, we, therefore, we propose a new, universally adaptable two-test system that could, in the case of SARS-CoV-2, perform better than any other known approach regardless of the actual seroprevalence: the sensitivity-improved Two-Test or SIT^2^. Its generalizability can be inferred from the following features: i) the adapted cut-offs used to optimize sensitivity were determined in various independent studies and were not explicitly calculated for our cohort (21-23); ii) SIT^2^ was effective, albeit with different efficiencies, in a total of 32 different test combinations; and iii) SIT^2^ was successfully validated in an independent cohort which was profoundly different from the derivation cohort. Therefore, SIT^2^ does not require a particular infrastructure or the availability of high-performance individual test systems but achieves the best performance out of an available test.

Our SIT^2^ strategy can rescue the specificity with minimal repeat testing required (see Table S6). For example, when applying the Roche NC as a screening test to our cohort, only 27 out of 1,181 samples needed confirmation testing with the Abbott NC test to correctly identify 62/64 true positives. Simultaneously, all false-positive results were eliminated, including those added by lowering the cut-offs (Table S4 and Fig. S1). Additionally, it was more sensitive than virus neutralization testing, which identified only 60/64 clinical positives (Fig. 3). This result is not completely surprising as it is known that not all patients who recovered from COVID-19 show detectable levels of neutralizing antibodies (29). Nevertheless, it should be noted that although antibody binding assays may have a higher sensitivity than neutralization assays, they only partially reflect the functional activity of SARS-CoV-2-reactive antibodies(30).

Our study has both strengths and limitations. One strength is the size of the cohorts examined, both in deriving the SIT^2^ algorithm (N=1,181) and validating it (N=1,512). The composition of our specificity cohort is also unique: it consists of three sub-cohorts with selection criteria to further challenge analytical specificity. The lower cut-offs used to increase sensitivity were not modeled within our datasets but were derived from ROC-analyses data of independent studies (21-23). Furthermore, we were able to test the performance of the two-test systems in a total of 32 combinations, 12 in the derivation cohort and another 20 combinations in the validation cohort. As a limitation, in the Austrian cohort, only samples ≥14 days after symptom onset were included. Therefore, no conclusions on the sensitivity of the early seroconversion phase can be made from these data. Furthermore, mild and asymptomatic cases were underrepresented in the British cohort, perhaps leading to an observed higher sensitivity of the test systems.

In conclusion, we describe the novel two-test algorithm SIT^2^, which makes it possible to maintain or even significantly improve sensitivity while approaching 100% specificity.

## Data Availability

Data is available for interested researchers upon request from the corresponding author.

## Abbreviations

CI: Confidence interval
COVID-19: Coronavirus disease 2019
ELISA: Enzyme-linked immunosorbent assay
ICU: Intensive Care Unit
NT: Neutralization titer
NC: Nucleocapsid
NP: Nucleoprotein
OTA: Orthogonal testing algorithms
PCR: Polymerase chain reaction
RBD: Receptor binding domain
SIT^2^: Sensitivity-Improved Two-Test
SARS-CoV-2: Severe Acute Respiratory Syndrome Coronavirus 2
S: Spike protein
VNT: Virus neutralization test

## Acknowledgments

We sincerely thank Marika Gerdov, Susanne Keim, Karin Mildner, Elisabeth Ponweiser, Manuela Repl, Ilse Steiner, Christine Thun, and Martina Trella for excellent technical assistance. Finally, we want to thank all the donors of the various study cohorts. The MedUni Wien Biobank is funded to participate in the biobank consortium BBMRI.at (www.bbmri.at) by the Austrian Federal Ministry of Science, Research and Technology. There was no external funding received for the work presented. However, test kits for the Technoclone ELISAs were kindly provided by the manufacturer. All authors have read the journal’s policy on disclosure of potential conflicts of interest and the following has to be disclosed: NP-N received a travel grant from DiaSorin. DWE reports lecture fees from Gilead outside the submitted work. OCB reports grants from GSK, grants from Menarini, grants from Boehringer Ingelheim, grants from Astra, grants from MSD, grants from Pfizer, and grants from Chiesi, outside the submitted work. SH does receive unrestricted research grants (GSK, Boehringer, Menarini, Chiesi, Astra Zeneca, MSD, Novartis, Air Liquide, Vivisol, Pfizer, TEVA) for the Ludwig Boltzmann Institute of COPD and Respiratory Epidemiology, and is on advisory boards for G. SK, Boehringer Ingelheim, Novartis, Menarini, Chiesi, Astra Zeneca, MSD, Roche, Abbvie, Takeda, and TEVA for respiratory oncology and COPD. PQ is an advisory board member for Roche Austria and reports personal fees from Takeda outside the submitted work. The Dept. of Laboratory Medicine (Head: OWF) received compensations for advertisement on scientific symposia from Roche, DiaSorin, and Abbott and holds a grant for evaluating an in-vitro diagnostic device from Roche. CJB is a Board Member of Technoclone. HH receives compensations for biobank services from Glock Health Science and Research and BlueSky immunotherapies.

## Notes

### Competing Interest Statement

NP received a travel grant from DiaSorin. DWE reports lecture fees from Gilead outside the submitted work. OCB reports grants from GSK, grants from Menarini, grants from Boehringer Ingelheim, grants from Astra, grants from MSD, grants from Pfizer, and grants from Chiesi, outside the submitted work. SH does receive unrestricted research grants (GSK, Boehringer, Menarini, Chiesi, Astra Zeneca, MSD, Novartis, Air Liquide, Vivisol, Pfizer, TEVA) for the Ludwig Boltzmann Institute of COPD and Respiratory Epidemiology, and is on advisory boards for G. SK, Boehringer Ingelheim, Novartis, Menarini, Chiesi, Astra Zeneca, MSD, Roche, Abbvie, Takeda, and TEVA for respiratory oncology and COPD. PQ is an advisory board member for Roche Austria and reports personal fees from Takeda outside the submitted work. The Dept. of Laboratory Medicine (Head: OWF) received compensations for advertisement on scientific symposia from Roche, DiaSorin, and Abbott and holds a grant for evaluating an in-vitro diagnostic device from Roche. CJB is a Board Member of Technoclone. HH receives compensations for biobank services from Glock Health Science and Research and BlueSky immunotherapies.

### Funding Statement

The MedUni Wien Biobank is funded for its participation in the biobank consortium BBMRI.at (www.bbmri.at) by the Austrian Federal Ministry of Science, Research and Technology. There was no external funding received for the work presented. However, test kits for the Technoclone ELISAs were kindly provided by the manufacturer free of charge.

### Author Declarations

Ethics committee of the Medical University of Vienna (1424/2020)

