## Supplementary material for "Increasing test specificity without impairing sensitivity – lessons learned from SARS-CoV-2 serology": S

### Supplemental File

#### Supplementary Methods

##### Evaluation cohorts (Vienna)

| | **SARS-CoV-2 Serology Specificity cohorts (all collected before 2020)** | | | **SARS-CoV-2 Serology Sensitivity Cohorts (COVID-19 cohort)** | | |
|---|---|---|---|---|---|---|
| **Cohort** | COHORT A<br><br>LEAD-study<br>(N=494) | COHORT B<br><br>MedUni Wien Biobank healthy donor collective<br>(N=265) | COHORT C<br><br>Cohort of rheumatic diseases<br>(N=359) | Post-COVID-19 donors at the MedUni Wien Biobank (N=38) | COVID-19-Convalescent plasma donors at the Department of Transfusion Medicine (N=7) | Diagnostic excess serum samples from COVID-19 patients sent to Department of Laboratory Medicine<br>(N=19) |
| **Cohort description** | Population-based cohort, representing a cross-section of the Viennese and surrounding area population; balanced for sex, age, and health status; collected before 2020 | Population- based cohort: samples from healthy donors collected at the MedUni Wien Biobank before 2020; | Disease-specific cohort: patients with a broad spectrum of rheumatic diseases from the Division of Rheumatology, Medical University of Vienna, collected before 2020 | Participants with a history of COVID-19, either confirmed by a positive SARS-CoV-2 RT-PCR (N=29) or symptomatic patients with close contact to PCR-positive COVID-19 patients (N=9) | Plasma donors with a history of SARS-CoV-2 RT-PCR confirmed COVID-19 | Samples from in- or outpatients with SARS-CoV-2 RT-PCR confirmed COVID-19<br><br>Intensive care unit patients (N=13) |
| **Inclusion criteria** | Age 6 – 80 years<br>Only material collected in the months November – April (enrichment of possible cross-reactive antibodies for seasonal respiratory infections) was used for this study | Age >18 years<br>Self-assessment: healthy | Age >18 years<br>Rheumatic disease | Age >18 years<br>positive SARS-CoV-2 RT-PCR OR close contact to patient with positive SARS-CoV-2 PCR considered healthy at the time of sample donation<br>written informed consent | Age >18 years<br>positive SARS-CoV-2 RT-PCR followed by 2 negative SARS-CoV-2 RT-PCRs<br>Considered healthy at the time of plasma donation<br>Written informed consent | Age >18 years<br><br>Positive SARS-CoV-2 RT-PCR |
| **Exclusion criteria** | Unavailability of biomaterial | Unavailability of biomaterial | Unavailability of biomaterial | | Inconclusive history of COVID-19<br>Unavailability of biomaterial | Unavailability of biomaterial |

**Table S1:** Cohort characteristics (Vienna)

| | LEAD-Study | Healthy donor collective | Rheumatic cohort |
|---|---|---|---|
| **Number (% of total)** | 494 (44%) | 265 (24%) | 358 (32%) |
| **Age (y)** | 43 (26 – 56) | 38 (25 – 52) | 52 (41 – 61) |
| **Female sex (% of cohort)** | 247 (50%) | 156 (59%) | 272 (76%) |
| **Samples collected (range)** | Dec 2011 – Apr 2016 | Feb 2012 – Dec 2019 | Feb 2006 – Mar 2017 |
| **Rheumatic disease (% of cohort)** | | | |
| *Rheumatoid Arthritis* | | | 142 (40%) |
| *Systemic lupus erythematosus* | | | 98 (27%) |
| *Systemic sclerosis* | | | 49 (14%) |
| *Psoriatic arthritis* | | | 25 (7%) |
| *Spondyloarthritis* | | | 25 (7%) |
| *Sjögren's Syndrome* | | | 19 (5%) |

**Table S2:** Demographical data of SARS-CoV-2 negative cohorts (Vienna). Unless otherwise stated, data are either presented as counts (percentages) or median (interquartile range).

| | COVID-19 cohort |
|---|---|
| **Number** | 64 |
| **Age (y)** | 49 (40 – 57) |
| **Female sex (% of cohort)** | 29 (45%) |
| **Symptom severity** | |
| *Asymptomatic* | 5 (8%) |
| *Mild* | 27 (42%) |
| *Moderate* | 15 (23%) |
| *Severe* | 4 (6%) |
| *Intensive care unit* | 13 (20%) |
| **Symptom onset (d before sampling)*** | 41 (26,25 – 49) |
| **Antibody response values** | |
| *Abbott [Index (S/C)], Cut-off 1·40* | 4·87 (2.77 – 6.85) |
| *Roche [COI], Cut-off 1·00* | 23·300 (7·220 – 52·900) |
| *DiaSorin [Au/ml], Cut-off 15·0 (borderline: 12-15)* | 48·1 (15·5 – 94·3) |

**Table S3:** Characteristics of SARS-CoV-2 positive individuals (Vienna). Data are either presented as counts (percentages) or median (interquartile range). *for asymptomatic individuals, PCR date was taken instead of symptom onset date.

##### SARS-CoV-2 antibody detection

The Roche Elecsys® Anti-SARS-CoV-2 assay detects total antibodies against the nucleocapsid (NC) antigen in a sandwich electrochemiluminescence immunoassay (ECLIA) on a cobas® e801 analyzer (Roche Diagnostics, Rotkreuz, Switzerland). According to the manufacturer, results exceeding 1·000 COI (cut-off index) are considered positive. The Abbott SARS-CoV-2 assay detects IgG-antibodies against NC in a chemiluminescence microparticle assay (CMIA) on Abbott ARCHITECT® i2000sr platforms (Abbott Laboratories, Chicago, USA). The cut-off suggested by the manufacturer is 1·40 index (S/C). The DiaSorin LIAISON® SARS-CoV-2 S1/S2 IgG test detects IgG-antibodies against the S1/S2 domains of the virus' spike protein on LIAISON® XL analyzers (DiaSorin S.p.A., Saluggia, Italy) employing chemiluminescence immunoassays (CLIA). Results >12·0 AU/mL are considered positive (the manufacturer suggests retesting of borderline results between 12·0 – 15·0 AU/mL; however, this approach is not practicable when using historical samples).

Commercially available quantitative ELISAs[1] are directed against the nucleoprotein (NP) (Technozym® anti SARS-CoV-2 NP IgG) or the RBD (Technozym® anti SARS-CoV-2 RBD IgG) domain of the spike protein (Technoclone, Vienna, Austria). Calibrators and controls of both tests are traceable to a specific antibody against the SARS-CoV-2 receptor-binding domain (CR3022). These quantitative assays were processed manually according to the manufacturer's instructions. IgG-Antibodies were quantified on a Filtermax F5 plate reader (Molecular Devices, San José, USA). The reduced cut-offs for these assays were modeled in-house and set at 3·000 U/ml for RBD and 3·500 U/ml for NP.

A serum neutralisation assay was used to determine SARS-CoV-2-specific neutralising antibodies against an authentic SARS-CoV-2 virus, originally isolated from a clinical specimen (B.1.414, Clade G, Spike D614G, GISAID accession ID EPI_ISL_583577). Assays were performed with Vero 76 clone E6 cells (CCLV-RIE929, Friedrich-Loeffler-Institute, Riems, Germany) cultured in minimum essential medium Eagle (E-MEM) with BioWhittaker Hank's balanced salt solution (HBSS) (Lonza, Basel, CH) supplemented with 10% (V/V) FBS (Corning Inc, Corning, NY). Neutralising antibody titers in heat-treated human sera were determined as previously described [1]. The cytopathic effect was evaluated and scored for each well using an inverted optical microscope. To determine neutralization titer the reciprocal of the highest serum dilution that protected more than 50% of the cells from the cytopathic effect was used and was calculated according to Reed and Muench[2].

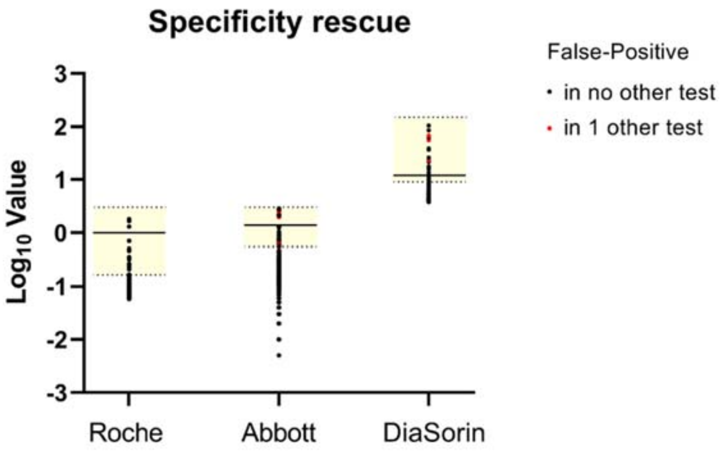


**Fig. S1:** Rescuing specificity by applying a confirmation test is possible, as false-positives (dots within yellow areas, which are limited by the adapted cut-offs for positivity [dotted lines]; the continuous line represents the manufacturer's cut-off) usually are negative in other SARS-CoV-2 antibody tests, with only a few exceptions (red dots).

**Supplementary Results**

| | | Roche | | Abbott | | DiaSorin | |
|---|---|---|---|---|---|---|---|
| | | *Manufacturer's cut-off:* | *Modified cut-offs:* | *Manufacturer's cut-off:* | *Modified cut-offs:* | *Manufacturer's cut-off:* | *Modified cut-offs:* |
| **A** | | **≥1·000 COI** | **>0·165 COI** | **≥1·40 Index** | **≥0·55 Index** | **≥12·0 AU/mL** | **≥9·0 AU/mL** |
| | Sensitivity | *89·1 (79·1-94·6)* | *98·4 (91·7-99·7)* | *84·4 (73·6-91·3)* | *96·9 (89·3-99·1)* | *82·8 (71·8-90·1)* | *89·1 (79·1-94·6)* |
| | Δ Sensitivity | | *Z=+2·45, P=0·014* | | *Z=+2·83, p=0·005* | | *Z=+2·00, p=0·046* |
| | Specificity | *99·7 (99·2-99·9)* | *98·4 (97·5-99·0)* | *99·2 (98·5-99·6)* | *97·6 (96·5-98·3)* | *98·2 (97·3-98·8)* | *97·2 (96·1-98·0)* |
| | Δ Specificity | | *Z=-3·87, P<0·001* | | *Z=-4·24, p<0·0001* | | *Z=-3·32, p<0·001* |
| **B** | | | **>3·000 COI** | | **>3·00 Index** | | **150·0 AU/mL** |
| | True positives | *57* | *54 (95% of TP)* | *54* | *45 (83% of TP)* | *53* | *5 (9% of TP)* |
| | False positives | *3* | *0* | *9* | *0* | *20* | *0* |

**Table S4.** Step 1 – Increasing sensitivity by cut-off adaption. A) Comparison of sensitivity, specificity and predictive values between results calculated using the cut-offs suggested by the manufacturers and reduced cut-offs derived from the literature. Green color indicates a significant gain, red color a significant reduction. B) Presentation of the proportion of true-positives (TP) that were excluded from re-testing by applying high cut-offs, which were determined after observations in our cohorts. Δ… Difference.

| **Roche** | **Manufacturer's cut-off:** | **Orthogonal testing with manufacturer's cut-offs:** | | | |
|---|---|---|---|---|---|
| **Cut-offs** | **≥1·000 COI** | **>1·000 COI + RBD >8 U/mL** | **>1·000 COI + NP >8 U/mL** | **>1·000 COI + Abbott ≥1·4 Index** | **>1·000 COI + DiaSorin ≥12·0 AU/mL** |
| Sensitivity | *89·1 (79·1-94·6)* | *81·3 (70·0-88·9)* | *78·1 (66·6-86·5)* | *84·4 (73·6-91·3)* | *79·7 (68·3-87·7)* |
| | | *Z=-2·24 P=0·025* | *Z=-2·65 P=0·008* | *Z=-1·73 P=0·083* | *Z=-2·45 P=0·014* |
| Specificity | *99·7 (99·2-99·9)* | *100·0 (99·7-100·0)* | *100·0 (99·7-100·0)* | *100·0 (99·7-100·0)* | *100·0 (99·7-100·0)* |
| | | *Z=+1·73 P=0·083* | *Z=+1·73 P=0·083* | *Z=+1·73 P=0·083* | *Z=+1·73 P=0·083* |

| **Abbott** | **Manufacturer's cut-off:** | **Orthogonal testing with manufacturer's cut-offs:** | | | |
|---|---|---|---|---|---|
| **Cut-offs** | **≥1·40 Index** | **≥1·40 Index + RBD >8 U/mL** | **≥1·40 Index + NP >8 U/mL** | **≥1·40 Index + Roche >1·000 COI** | **≥1·40 Index + DiaSorin ≥12·0 AU/mL** |
| Sensitivity | *84·4 (73·6-91·3)* | *78·1 (66·6-86·5)* | *78·1 (66·6-86·5)* | *84·4 (73·6-91·3)* | *75·0 (63·2-84·0)* |
| | | *Z=-2·00 P=0·046* | *Z=-2·00 P=0·046* | *Z=±0·00* | *Z=-2·45 P=0·014* |
| Specificity | *99·2 (98·5-99·6)* | *100·0 (99·7-100·0)* | *99·9 (99·5-100·0)* | *100·0 (99·7-100·0)* | *99·9 (99·5-100·0)* |
| | | *Z=+3·00 P=-0·003* | *Z=+2·83 P=0·005* | *Z=+3·00 P=-0·003* | *Z=+2·83 P=0·005* |

| **DiaSorin** | **Manufacturer's cut-off:** | **Orthogonal testing with manufacturer's cut-offs:** | | | |
|---|---|---|---|---|---|
| **Cut-offs** | **≥12·0 AU/mL*** | **≥12·0 AU/mL + RBD >8 U/mL** | **≥12·0 AU/mL + NP >8 U/mL** | **≥12·0 AU/mL + Roche >1·000 COI** | **≥12·0 AU/mL + Abbott ≥1·4 Index** |
| Sensitivity | *82·8 (71·8-90·1)* | *76·6(64·9-85·3)* | *70·3 (58·2-80·1)* | *79·7 (68·3-87·7)* | *75·0 (63·2-84·0)* |
| | | *Z=-2·00 P=0·046* | *Z=-2·83 P=0·005* | *Z=-1·41 P=0·157* | *Z=-2·24 P=0·025* |
| Specificity | *98·2 (97·3-98·8)* | *99·9 (99·5-100·0)* | *100·0 (99·7-100·0)* | *100·0 (99·7-100·0)* | *99·9 (99·5-100·0)* |
| | | *Z=+4·36 P<0·0001* | *Z=+4·47 P<0·0001* | *Z=+4·47 P<0·0001* | *Z=+4·36 P<0·0001* |

**Table S5.** Effect of an orthogonal test strategy (OTA) with the tests used as suggested by the manufacturer. Resulting sensitivities and specificities are compared to those yielded if the respective single test is used as suggested by the manufacturer. Green color indicates a significant gain, yellow color a specificity of approximately 100·0% that could not be further improved, and red color a significant loss. N… Number; Δ… Difference; *… Values between 12 – 15 AU/ml should be considered borderline and require re-testing at a later time-point, which is not possible in a cross-sectional setting. Therefore, these results were treated as positive.

| **Roche** | **Manufacturer's cut-off:** | **Sensitivity-improved Two-Test Approach:** | | | |
|---|---|---|---|---|---|
| **Cut-offs** | **≥1·000 COI** | **≤0·165 COI = 0; ≥3 COI = 1;<br>>0·165 AND <3·000 COI = 1 if:** | | | |
| | | **+ RBD >3·0 U/mL** | **+ NP >3·5 U/mL** | **+ Abbott ≥0·55 Index** | **+ DiaSorin ≥9·0 AU/mL** |
| N (%) retested | | *27 (2·3%)* | | | |
| Sensitivity | *89·1 (79·1-94·6)* | *95·3 (87·1-98·4)* | *92·2 (83·0-96·6)* | *96·9 (89·3-99·1)* | *95·3 (87·1-98·4)* |
| Δ Sensitivity | | *Z=+2·00, P=0·046* | *Z=+1·00, P=0·317* | *Z=+2·24, P=0·025* | *Z=+2·00, P=0·046* |
| Specificity | *99·7 (99·2-99·9)* | *100·0 (99·7-100·0)* | *100·0 (99·7-100·0)* | *100·0 (99·7-100·0)* | *100·0 (99·7-100·0)* |
| Δ Specificity | | *Z=+1·73, P=0·083* | *Z=+1·73, P=0·083* | *Z=+1·73, P=0·083* | *Z=+1·73, P=0·083* |

| **Abbott** | **Manufacturer's cut-off:** | **Sensitivity-improved Two-Test Approach:** | | | |
|---|---|---|---|---|---|
| **Cut-offs** | **≥1·40 Index** | **<0·55 Index = 0; ≥3 Index = 1;<br>≥0·55 AND <3·000 Index = 1 if:** | | | |
| | | **+ RBD >3·0 U/mL** | **+ NP >3·5 U/mL** | **+ Roche >0·165 COI** | **+ DiaSorin ≥9·0 AU/mL** |
| N (%) retested | | *45 (3·8%)* | | | |
| Sensitivity | *84·4 (73·6-91·3)* | *95·3 (87·1-98·4)* | *92·2 (83·0-96·6)* | *96·9 (89·3-99·1)* | *92·2 (83·0-96·6)* |
| Δ Sensitivity | | *Z=+2·65, p=0·008* | *Z=+2·24, p=0·025* | *Z=+2·83, p=0·005* | *Z=+1·89, p=0·059* |
| Specificity | *99·2 (98·5-99·6)* | *99·8 (99·3-100·0)* | *99·8 (99·3-100·0)* | *100·0 (99·7-100·0)* | *99·9 (99·5-100·0)* |
| Δ Specificity | | *Z=+2·33, p=0·020* | *Z=+2·13, p=0·020* | *Z=+3·00, p=0·003* | *Z=+2·83, p=0·005* |

| **DiaSorin** | **Manufacturer's cut-off:** | **Sensitivity-improved Two-Test Approach:** | | | |
|---|---|---|---|---|---|
| **Cut-offs** | **≥12·0 AU/mL** | **<9·0 AU/mL = 0; ≥150 AU/mL = 1;<br>≥9·0 AND <150·0 AU/mL = 1 if:** | | | |
| | | **+ RBD >3·0 U/mL** | **+ NP >3·5 U/mL** | **+ Roche >0·165 COI** | **+ Abbott ≥0·55 Index** |
| N (%) retested | | *87 (7·4%)* | | | |
| Sensitivity | *82·8 (71·8-90·1)* | *87·5 (77·2-93·5)* | *84·4 (73·6-91·3)* | *89·1 (79·1-94·6)* | *87·5 (77·2-93·5)* |
| Δ Sensitivity | | *Z=+1·34, p=0·180* | *Z=+0·38, p=0·701* | *Z=+2·00, p=0·046* | *Z=+1·34, p=0·180* |
| Specificity | *98·2 (97·3-98·8)* | *99·8 (99·3-100·0)* | *100·0 (99·7-100·0)* | *100·0 (99·7-100·0)* | *99·9 (99·5-100·0)* |
| Δ Specificity | | *Z=+4·24, p<0·0001* | *Z=+4·47, p<0·0001* | *Z=+4·47, p<0·0001* | *Z=+4·36, p<0·0001* |

**Table S6.** Effect of a sensitivity-improved two-test approach ($SIT^2$). Resulting sensitivities and specificities are compared to those yielded if the respective single test is used as suggested by the manufacturer. N… Number; Δ… Difference.

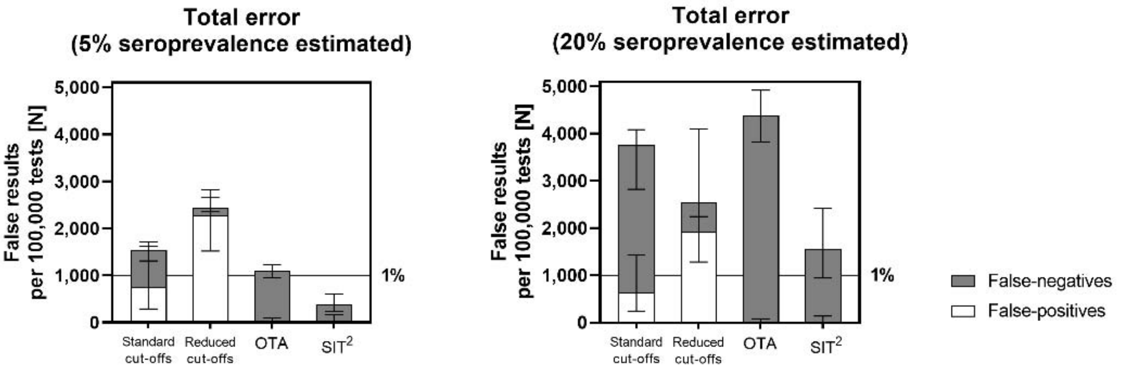


**Fig. S2:** Median and interquartile ranges of total error, subdivided by error produced by false-positivity (white) or false-negativity (grey). OTA… orthogonal testing algorithms; SIT$^2$… Sensitivity improved two-test; $N_{\text{Standard cut-offs}}$ = 3, $N_{\text{Reduced cut-offs}}$ = 3, $N_{\text{OTA}}$, $N_{\text{SIT}^2}$ = 12.

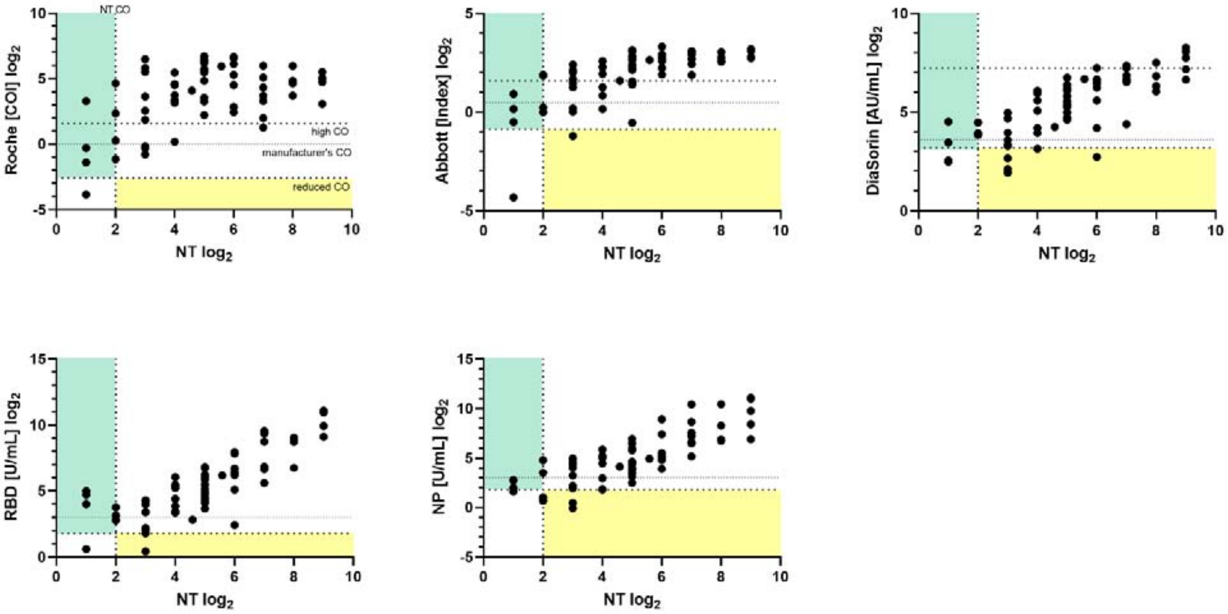


**Fig. S3:** Relationship between antibody levels detected by binding assays (Roche, Abbott, DiaSorin, Technozym RBD and NP) and viral neutralization test (TCID50) titers in 64 individuals after SARS-CoV-2 infection. Sera within the green area are positive in a sensitivity improved binding assay, but yield a negative TCID50. Within the yellow area, sera present with a positive TCID50, whereas binding assay results remain negative after sensitivity improvement. CO… cut-off, NT… viral neutralization test.

#### Figure data

| A Single tests | | | | | | | | |
|---|---|---|---|---|---|---|---|---|
| **Test** | **Sens [%]** | **Spec [%]** | **FP 5%** | **FP20%** | **FN5%** | **FN20%** | **TE5%** | **TE20%** |
| **Roche MC** | *89·1* | *99·7* | *285* | *240* | *545* | *2180* | *830* | *2420* |
| **Abbott MC** | *84·4* | *99·2* | *760* | *640* | *780* | *3120* | *1540* | *3760* |
| **DiaSorin MC** | *82·8* | *98·2* | *1710* | *1440* | *860* | *3440* | *2570* | *4880* |
| **Roche RC** | *98·4* | *98·4* | *1520* | *1280* | *80* | *320* | *1600* | *1600* |
| **Abbott RC** | *96·9* | *97·6* | *2280* | *1920* | *155* | *620* | *2435* | *2540* |
| **DiaSorin RC** | *89·1* | *97·2* | *2660* | *2240* | *545* | *2180* | *3205* | *4420* |

**B1 OTA False-positives (5%/20% seroprevalence) per 100,000 tests**

| Confirmation / Screening | Roche | Abbott | DiaSorin | RBD | NP |
|---|---|---|---|---|---|
| **Roche** | | *0/0* | *0/0* | *0/0* | *0/0* |
| **Abbott** | *0/0* | | *95/80* | *0/0* | *95/80* |
| **DiaSorin** | *0/0* | *95/80* | | *95/80* | *0/0* |

**B2 OTA False-negatives (5%/20% seroprevalence) per 100,000 tests**

| Confirmation / Screening | Roche | Abbott | DiaSorin | RBD | NP |
|---|---|---|---|---|---|
| **Roche** | | *780/3120* | *1015/4060* | *935/3740* | *1095/4380* |
| **Abbott** | *780/3120* | | *1250/5000* | *1095/4380* | *1095/4380* |
| **DiaSorin** | *1015/4060* | *1250/5000* | | *1170/4680* | *1485/5940* |

**B3 OTA Total error (5%/20% seroprevalence) per 100,000 tests**

| Confirmation / Screening | Roche | Abbott | DiaSorin | RBD | NP |
|---|---|---|---|---|---|
| **Roche** | | *780/3120* | *1015/4060* | *935/3740* | *1095/4380* |
| **Abbott** | *780/3120* | | *1345/5080* | *1095/4380* | *1190/4460* |
| **DiaSorin** | *1015/4060* | *1345/5080* | | *1265/4760* | *1485/5940* |

**C1 SIT² False-positives (5%/20% seroprevalence) per 100,000 tests**

| Confirmation / Screening | Roche | Abbott | DiaSorin | RBD | NP |
|---|---|---|---|---|---|
| **Roche** | | *0/0* | *0/0* | *0/0* | *0/0* |
| **Abbott** | *0/0* | | *85/80* | *190/160* | *190/160* |
| **DiaSorin** | *0/0* | *95/80* | | *190/160* | *0/0* |

**C2 SIT² False-negatives (5%/20% seroprevalence) per 100,000 tests**

| Confirmation / Screening | Roche | Abbott | DiaSorin | RBD | NP |
|---|---|---|---|---|---|
| **Roche** | | *155/620* | *235/940* | *235/940* | *390/1560* |
| **Abbott** | *155/620* | | *390/1560* | *235/940* | *390/1560* |
| **DiaSorin** | *545/2180* | *625/2500* | | *625/2500* | *780/3120* |

**C3 SIT² Total error (5%/20% seroprevalence) per 100,000 tests**

| Confirmation / Screening | Roche | Abbott | DiaSorin | RBD | NP |
|---|---|---|---|---|---|
| **Roche** | | *155/620* | *235/940* | *235/940* | *390/1560* |
| **Abbott** | *155/620* | | *485/1640* | *425/1100* | *580/1720* |
| **DiaSorin** | *545/2180* | *720/2580* | | *815/2660* | *780/3120* |

**Figure data of Fig. 2:** A) Quality criteria (Sens… Sensitivity, Spec… Specificity, FP… false-positives, FN… false-negatives, TE… total error), whenever applicable estimated for 5% or 20% seroprevalence. MC… manufacturer's cut-off, RC… reduced cut-off according to published data (>0·165 COI for Roche, ≥0·55 Index for Abbott, ≥9·0 AU/mL for DiaSorin). B1-3) False-positives, false-negatives and total error, each at 5 or 20% estimated seroprevalence, for different Orthogonal Testing Algorithm (OTA) combinations, where positives of the screening test (vertical) have to be confirmed by a confirmation test (horizontal). C1-3) False-positives, false-negatives and total error, each at 5 or 20% estimated seroprevalence, for different Sensitivity-improved Two-Test (SIT²) combinations.
